# Discordance Between Genetic Ancestry and Self-Reported Race Impacts Inference of Neuropsychiatric Burden in Alzheimer’s Disease

**DOI:** 10.64898/2026.08.23.26361161

**Authors:** Ajneesh Kumar, Balaji Kannappan, Nicholas R. Ray, Jiji T. Kurup, Pamela Del Rosario, Alyssa N. De Vito, Michael L. Cuccaro, Gary W. Beecham, Edward D. Huey, Christiane Reitz

## Abstract

**Introduction:** Neuropsychiatric symptoms (NPS)—including aggression, psychosis, anxiety, apathy, and depression—affect up to 85% of individuals with Alzheimer’s disease (AD) and are among its most disabling and costly manifestations, accelerating cognitive and functional decline, institutionalization, mortality, and healthcare costs. NPS prevalence has largely been characterized using self-reported race. Whether NPS differs across genetically defined ancestry groups—and whether self-reported race obscures these differences—remains unknown, limiting accurate risk stratification and treatment development.

**Methods:** Using whole-genome sequencing data from 7,118 ADSP participants, we defined three NPS clusters from the NPI-Q: early psychosis (CDR 0.5–1), late psychosis (CDR 2–3), and affective symptoms. Genetic ancestry was inferred by principal component clustering, identifying six groups (EUR, AFR, EAS, SAS, AMR, ADMIXED), and compared with self-reported race/ethnicity. NPS prevalence was compared across genetic ancestry groups and genetic ancestry and self-reported race using Fisher’s exact and regression models.

**Results:** Genetic ancestry assignment differed markedly from self-reported race, affecting NPS prevalence estimates. NPS prevalence also differed across ancestry groups; affective symptoms were highest in EAS (90%) and SAS (77%) and lowest in AFR (66%), while psychosis was highest in EAS (74%) and SAS (70%) and lowest in AMR (55%) and EUR (56%), with similar patterns for early and late psychosis.

## Discussion

Genetically defined ancestry alters NPS prevalence estimates in AD, suggesting that standard race categories obscure population-level disease burden and compromise risk stratification, screening, and trial design. Ancestry-associated differences suggest partially distinct genetic and environmental drivers, underscoring the need to incorporate genetic ancestry into AD research and care.

## INTRODUCTION

Neuropsychiatric symptoms (NPS)—including depression, anxiety, apathy, agitation, psychosis, sleep disturbance, and appetite changes—are core clinical features of Alzheimer’s disease (AD), affecting over 85% of AD cases over the disease course.[1] NPS are associated with faster cognitive and functional decline, earlier institutionalization, increased caregiver burden, and higher mortality.[1] Despite their major impact, the biological factors contributing to variability in NPS expression remain poorly understood, limiting the development of targeted, mechanism-based interventions.

Population differences in NPS prevalence have traditionally been examined using self-identified race and ethnicity.[2, 3] While these measures are essential for understanding the influence of lived experience and structural inequities, they do not capture the underlying genetic architecture accurately [4]. As a result, studies relying solely on self-reported race may conflate social, environmental, and biological influences, hindering identification of the biological pathways that contribute to neuropsychiatric heterogeneity in AD.[5] Advancing precision medicine requires approaches that more directly reflect inherited variation relevant to disease mechanisms.

Genetically inferred ancestry, derived from genome-wide data, provides a complementary and biologically informative framework for characterizing population structure.[5] In diverse and admixed populations, genetic ancestry captures continuous patterns of shared genetic variation that are not represented by discrete racial categories and may better align with differences in disease-relevant mechanistic pathways.[5, 6] Examining NPS across genetically defined ancestry groups therefore offers an opportunity to refine phenotypes and their impact on disease burden, improve gene–symptom mapping, and generate hypotheses about ancestry-linked biological mechanisms, while facilitating modeling social determinants of health explicitly rather than embedding them within racial labels.

In the present study we compared NPS prevalence in AD using self-reported race and genetically inferred ancestry, directly evaluating differences across ancestry groups. This approach advances a biologically informed, precision-medicine framework for NPS while providing critical public health insights for identifying high-risk populations and guiding targeted screening, resources, and interventions.

## METHODS

### Study population

Included in the study were 7,118 individuals with AD from diverse ancestries from the Alzheimer’s Disease Sequencing Project (ADSP; https://adsp.niagads.org/).

Demographic information, including age, sex, and education, was collected using standardized protocols. Self-reported race was defined according to U.S. federal standards[7] and categorized as White, Black or African American, Asian, Native American/Alaska Native, Native Hawaiian/Other Pacific Islander, or more than one race.

### Cognitive and Neuropsychiatric data

In all contributing studies, participants have completed standard clinical assessments using established criteria for the diagnosis of AD.[8–10] Domain specific impairments in activities of daily living, cognition, and NPS are assessed. Assessments include a participant interview, neurological examination, and a Clinical Dementia Rating (CDR®)[11] to assign degree of impairment. NPS were assessed using the Neuropsychiatric Interview Questionnaire (NPI-Q),[12] which is an informant-based assessment of the severity of 12 NPS common in AD adapted from the full NPI.[13] Each NPS is coded as absent or present by a rater and if present is given a severity rating from 1 (mild) to 3 (severe). The NPI-Q is one of the most commonly used measures of NPS in AD.[14] It has been cross-validated with the NPI and shown good test-retest reliability.[13] The NPI-Q has been translated into over 10 languages and shown good reliability across translations.[15] Cultural differences in the reporting of NPS may exist but are difficult to distinguish from biological effects and the influences of other social determinants of health [16]. Comparisons of patients of different ancestry groups on the NPI-Q show a similar factor structure of the NPI-Q and differences on the total NPI-Q and individual NPS.[2, 17]

To refine NPS phenotypes we created the following 4 symptom clusters: Early and late psychosis and early and late affective symptoms. Participants with the psychosis phenotype had either delusions or hallucinations of any severity as assessed on the NPI Q at any visit. Timing of psychotic symptoms was defined based on when the symptoms occurred during the stage of the illness: early psychosis (EPS) is defined as the initial presence of psychotic symptoms at a CDR of 0.5 or 1. Late psychosis (LPS) is defined as the initial presence of psychotic symptoms at a CDR of 2 or 3. This distinction is based on previous studies demonstrating that the disease course in patients with early psychosis differs from patient’s without early psychosis [18]. Participants classified as having neither LPS nor EPS (PS–) do not meet the above psychotic symptoms criteria at any visit. The affective phenotype is defined by presence of depression, anxiety, and/or irritability of any severity on the NPI Q at any visit (AS+). This phenotype is based on observations that depression, anxiety, and irritability are often co-linear [19]. Timing of AS is defined based on initial presentation of symptoms: early affective symptoms (EAS) is defined as the initial presence of affective symptoms at CDR of 0.5 or 1, with late affective symptoms (LAS) at a CDR of 2 or 3; those classified as having neither (AS–) do not meet the above criteria at any visit.[20]

### Genetic Ancestry Estimation

Genetic ancestry was derived from ADSP R5 whole-genome sequencing data, using the full ADSP sample of 55,903 individuals. Harmonization of genomic data is performed by the Genome Center for Alzheimer’s Disease (GCAD; U54AG052427; PI: Wang) utilizing the VCPA[21] pipeline developed for the ADSP.. The approach uses GATK[22, 23] for SNV/Indel calling. The workflow includes mapping reads to hg38, BAM sorting, duplicate marking, quality scores, and local read realignments around known indels. GATK HaplotypeCaller is then applied to generate individual genotype calls in genomic and project-level VCF formats. Variant-level quality metrics include VQSR quality tranches, call rates, average read depths, excessive read depths [>500 reads], and excess heterozygosity or departure from HWE. Sample-level QC includes within-sample genotype call rate, Ti/Tv ratio for SNVs, heterozygosity/homozygosity ratio, and excess burden of singleton/doubleton variants. Larger structural variants (SVs) are called by the ADSP-SV working group, which uses multiple calling pipelines to generate a high-quality set of SVs from the ADSP WGS data. To estimate genetic ancestry. principal component analysis (PCA) was applied to capture population structure, followed by K-means clustering on the first ten principal components and ADMIXTURE (v0.03)[24] to define discrete genetically inferred ancestry groups. The resulting clusters represented major continental and admixed ancestry groups within the cohort.

### Statistical Analyses

We compared NPS prevalence between groups defined by self-reported race and genetically inferred ancestry, as well as across genetically defined ancestry groups. Cohen’s kappa statistics were calculated to assess the level of agreement between classifications beyond that expected by chance. Fisher’s exact test was used for initial pairwise comparisons of categorical symptom prevalence. Multivariable logistic regression models were subsequently applied to evaluate associations while adjusting for age, sex, and education. Odds ratios (ORs) and 95% confidence intervals (CIs) were reported to quantify differences in NPS prevalence across both self-reported race and genetic ancestry groups. All analyses were conducted using R (version 4.4.0) and significance was set at a two-sided p-value <0.05.

## RESULTS

### Categorization of individuals based on self-reported race and genetic ancestry

Table 1 shows the characteristics of the 7,118 individuals in the study sample. Based on self-reported race/ethnicity according to federal standards[7], 4451 individuals were non-Hispanic White, 688 were Hispanic White, 257 reported to be White without further specification, 805 identified as Black or African American, 75 identified as Asian, 46 identified as American Indian/Alaska Native, and 796 were grouped as “other” or “unknown”. Principal component analysis (PCA) identified six genetic ancestry clusters: European (EUR), African (AFR), East Asian (EAS), South Asian (SAS), Amerindian (AMR), and admixed (ADMIXED) (Figure 1). Of the 7,118 individuals with NPS information included in the present analyses, 4,870 clustered with EUR ancestry, 1,193 ADMIXED, 689 AFR, 59 EAS, 183 AMR, and 124 SAS.

**Figure 1.**
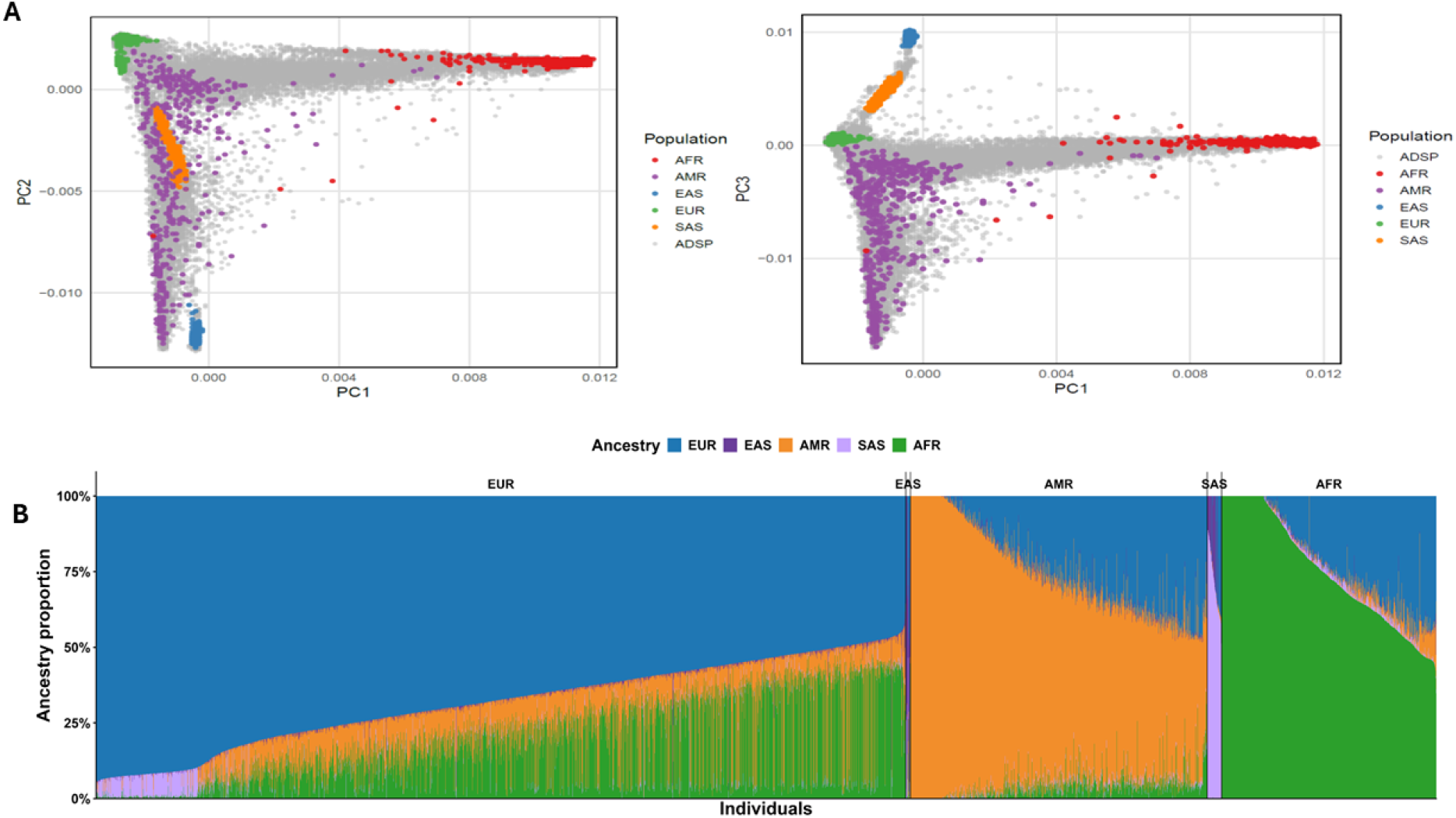
Genetic Ancestry Clusters, (A) Genomic PCA with ADSP participants shown in gray and global reference population samples color-coded as shown in the key. Left panels show PC1 versus PC2 comparisons, and right panels show PC1 versus PC3 comparisons, with the percent of variance explained by each PC shown. (B) Genetic ancestry proportions for ADSP participants stratified by the genetic similarity groups shown in Figure 1A.

**Table 1.** Demographic Characteristics of the Study Sample by self-reported Race and Genetic Ancestry Clusters.

| **Ethnicity/Genetic Ancestry** | **N** | **Age**<br>**(Mean ± SD)** | **Female, n (%)** |
| --- | --- | --- | --- |
| Overall | 7,118 | 71.4 ± 10.9 | 4,166 (58.5%) |
| **Categorized by Self-reported Race** |  |  |  |
| Non-Hispanic White | 4,451 | 70.7 ± 11.3 | 2,434 (54.7%) |
| Hispanic White | 688 | 73.4 ± 9.6 | 441 (64.1%) |
| Unspecified White | 257 | 68.8 ± 9.7 | 138 (53.7%) |
| Black or African American | 805 | 74.3 ± 9.5 | 565 (70.2%) |
| Asian | 75 | 65.2 ± 9.3 | 44 (58.7%) |
| American Indian/Alaska Native | 46 | 67.7 ± 9.9 | 24 (52.2%) |
| Other | 526 | 74.1 ± 10.6 | 362 (68.8%) |
| Unknown | 270 | 70.4 ± 10.5 | 158 (58.5%) |
| **Categorized by Genetic Ancestry** |  |  |  |
| EUR | 4,870 | 70.7 ± 11.1 | 2,675 (54.9%) |
| AFR | 689 | 74.8 ± 9.5 | 494 (71.7%) |
| EAS | 59 | 64.7 ± 9.3 | 35 (59.3%) |
| SAS | 124 | 69.4 ± 11.5 | 76 (61.3%) |
| ADMIXED | 1,193 | 73.6 ± 10.0 | 784 (65.7%) |
| AMR | 183 | 69.0 ± 10.1 | 102 (55.7%) |

Transitions between self-reported race/ethnicity and genetic ancestry clusters are shown in Figure 2 and Supplemental Table 1. While nearly all individuals self-identifying as non-Hispanic White (n=4,451) or unspecified White (n=257) clustered with European ancestry (EUR) forming the largest genetic ancestry group (n=4,870), genetic ancestry significantly diverged from self-reported ethnicity in all other groups. Among Hispanic White individuals, 68% mapped to ADMIXED ancestry, but 19.5% mapped to EUR, 11.5% to South Asian (SAS), and ∼1% to Amerindian (AMR) or African (AFR). Self-reported Black or African American individuals (n=805) mostly clustered with AFR (77%), but 23% mapped to ADMIXED and <1% to EUR or SAS. Among Asians (n=75), 73% mapped to East Asian (EAS), 13.4% to SAS, and 10.7% to ADMIXED. American Indian/Alaska Native individuals (n=46) showed mixed ancestry, with 41.3% ADMIXED, 37% EUR, and 17.4% AMR. High heterogeneity was also observed among those reporting “Other” (n=526), who mapped primarily to ADMIXED (70%), with smaller contributions from AFR (11%), AMR (11%), EUR (6%), and SAS (2%), and among those with missing self-report (“NA”, n=270), who mapped mainly to ADMIXED (49%) and AMR (41.5%), with 7% SAS. Agreement between self-reported race and genetically inferred ancestry varied across ancestry groups. Strong agreement was observed for Non-Hispanic White vs. European ancestry (κ = 0.86), Asian vs. East Asian ancestry (κ = 0.82), and Black or African American vs. African ancestry (κ = 0.81). In contrast, agreement was low for Asian vs. South Asian ancestry (κ = 0.09), American Indian/Alaska Native vs. American Indian ancestry (κ = 0.06), and American Indian/Alaska Native and Admixed ancestry (κ = 0.02). Hispanic White and European ancestry also showed disagreement (κ = −0.15) Supplementary Figure 1.

**Figure 2.**
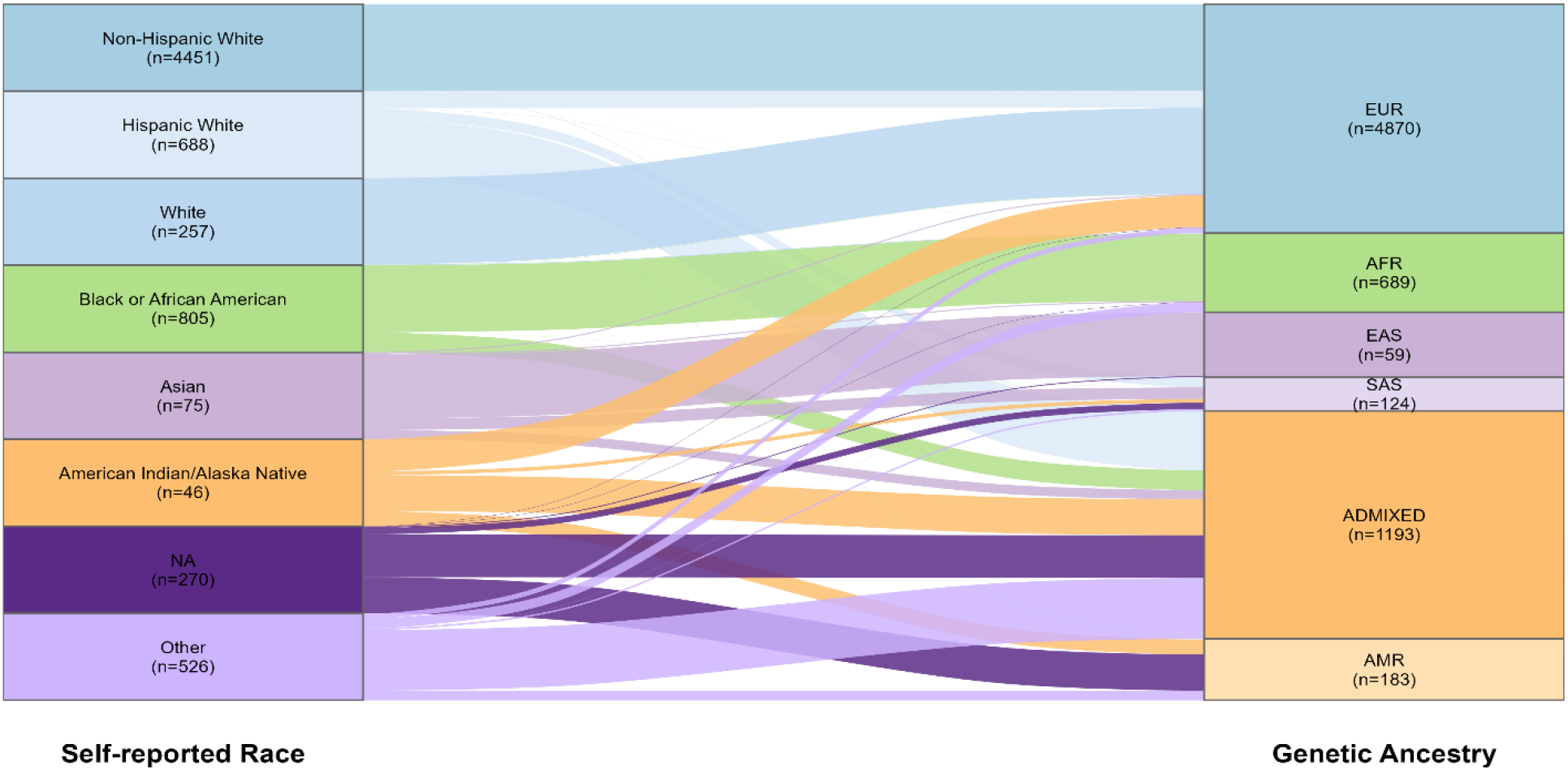
Sankey diagram illustrating the relationship between self-reported race/ethnicity and genetically inferred ancestry in the study sample. Each vertical column represents a set of participants defined by either self-reported race (left) or genetic ancestry cluster (right). Flows between columns indicate how individuals in each self-reported racial/ethnic category are assigned to genetic ancestry groups based on principal component analysis, K-means clustering and admixture analysis. The width of each flow is proportional to the number of participants, highlighting concordance and divergence between social and biological definitions of population structure.

### Comparison of NPS prevalence in AD between self-reported race and genetic ancestry groups, and across genetic ancestry groups

Comparison of NPS prevalence in AD between self-reported race and genetic ancestry showed individuals with self-reported Black ethnicity and AFR ancestry broadly concordant results for both affective symptoms (∼67%) and psychosis (∼65%) among (Figures 3 and 4; Supplemental Table 2). In contrast, discrepancies were observed between self-reported NHW or HW ethnicity versus EUR ancestry, self-reported Asian ethnicity versus SAS and EAS ancestry, and self-reported American Indian/Alaska Native versus AMR and ADMIXED ancestry. Notably, these differences were most pronounced and consistently statistically significant for affective symptoms, which showed for genetically defined EUR ancestry significantly lower prevalences compared to self-reported NHW ethnicity (EUR: 72%, NHW: 76%; p=1.45 × 10); for genetically defined ADMIXED (69%, p=0.008) and AMR ancestry (71%, p=0.03) significantly lower prevalences compared to self-reported American Indian/Alaska Native ethnicity (87%), and for genetically defined SAS ancestry a significantly lower prevalence compared to self-reported Asian ethnicity (SAS: 77%; Asian: 88%, p=0.08) that approached statistical significance. Psychosis prevalences, and differences in psychosis prevalence (overall, early, and late), were generally smaller in magnitude and only statistically significant in EUR ancestry vs. self-reported non-Hispanic White ethnicity (EUR: 56%; NHW: 59%; p=0.002). When examining NPS prevalence across genetic ancestry clusters, affective symptoms were highest in EAS (90%) and SAS (77%) and lowest in AFR (66%) (Figure 7). In contrast, psychosis prevalence, largely driven by early psychosis, was consistently lower across all ancestry groups, with the highest rates again observed in EAS (74%) and SAS (70%), but the lowest rates occurring in EUR (56%) and AMR (55%), rather than AFR.

**Figure 3.**
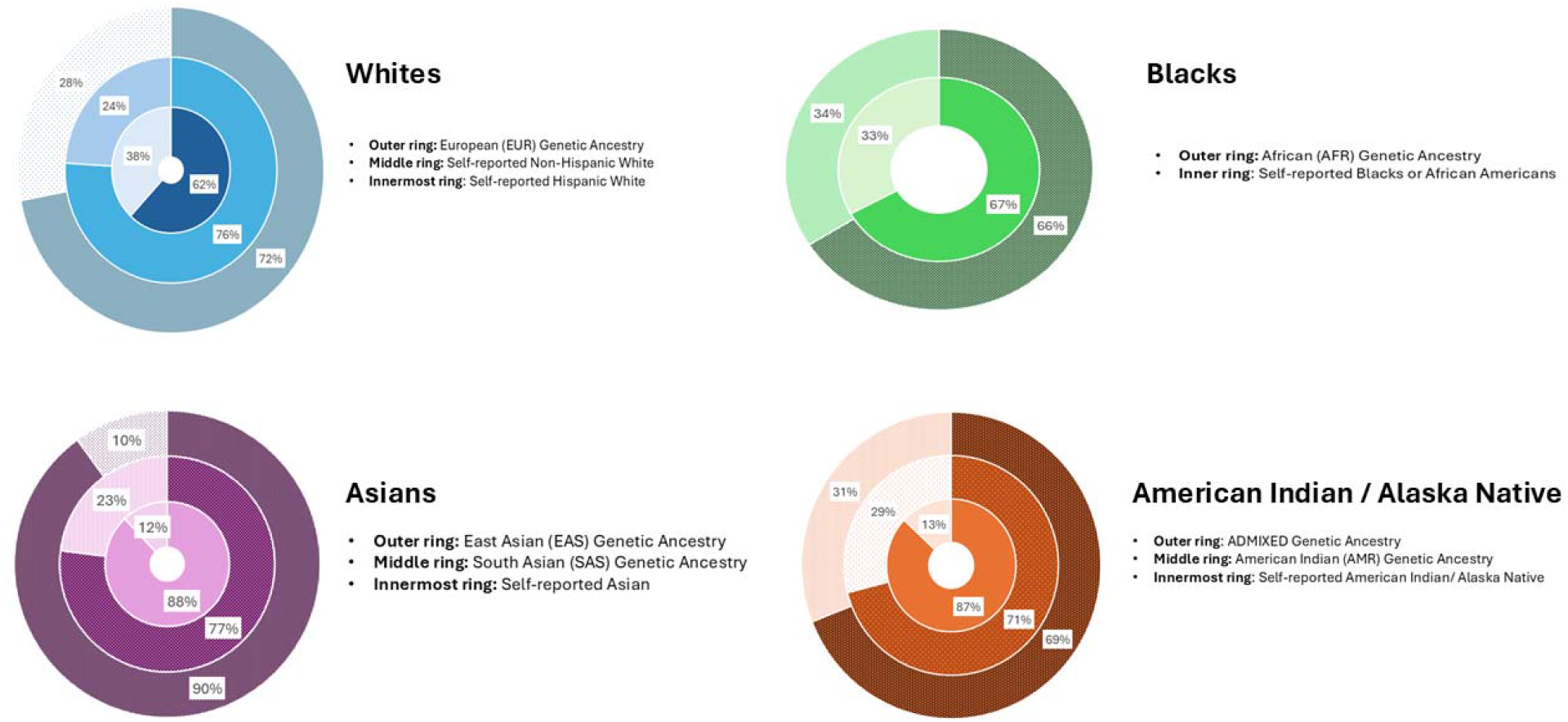
Prevalence of Affective Symptoms in AD when using self-reported ancestry vs. genetic ancestry. For each ring, the darker color represents the prevalence of having the symptom, while the lighter color represents the prevalence of not having the symptom.

**Figure 4.**
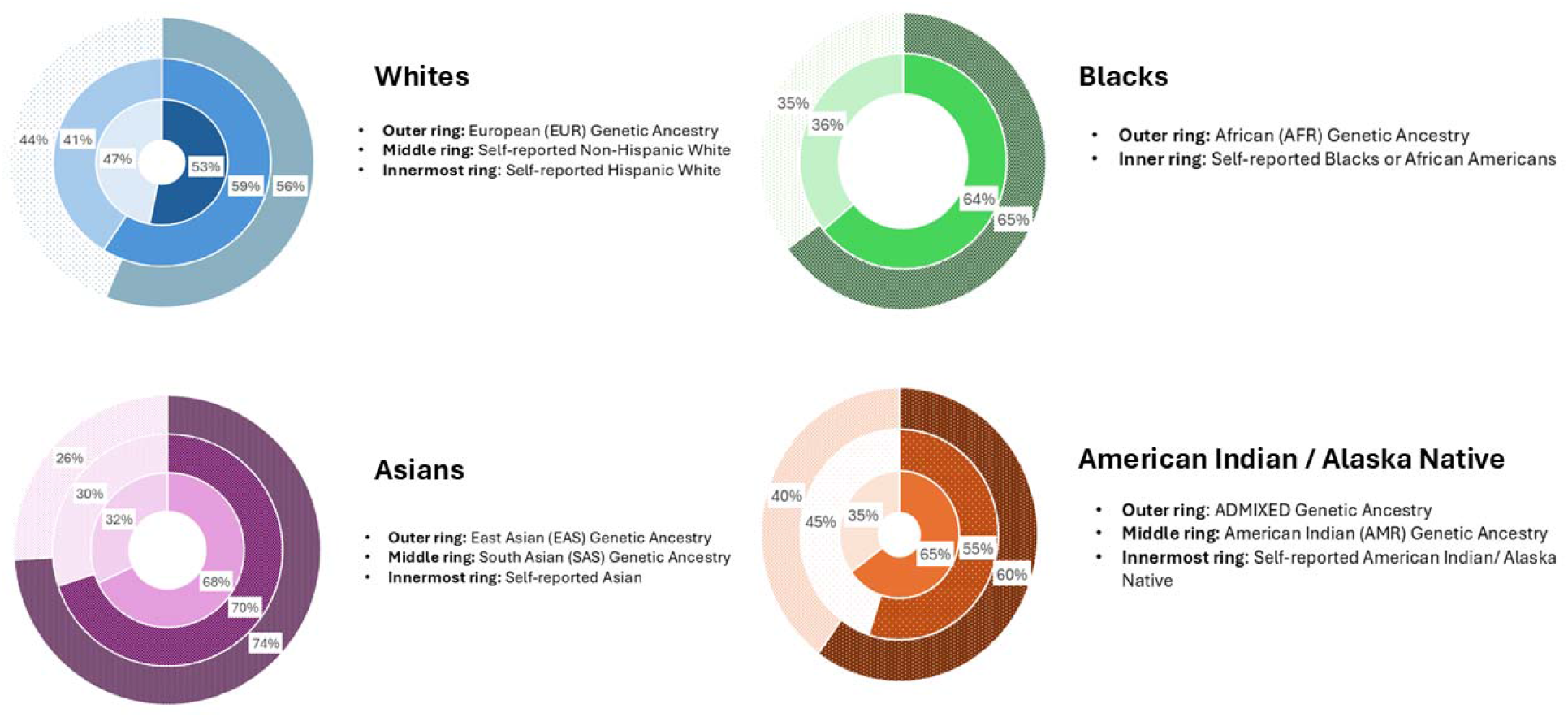
Prevalence of Psychosis in AD when using self-reported ancestry vs. genetic ancestry. For each ring, the darker color represents the prevalence of having the symptom, while the lighter color represents the prevalence of not having the symptom.

**Figure 5.**
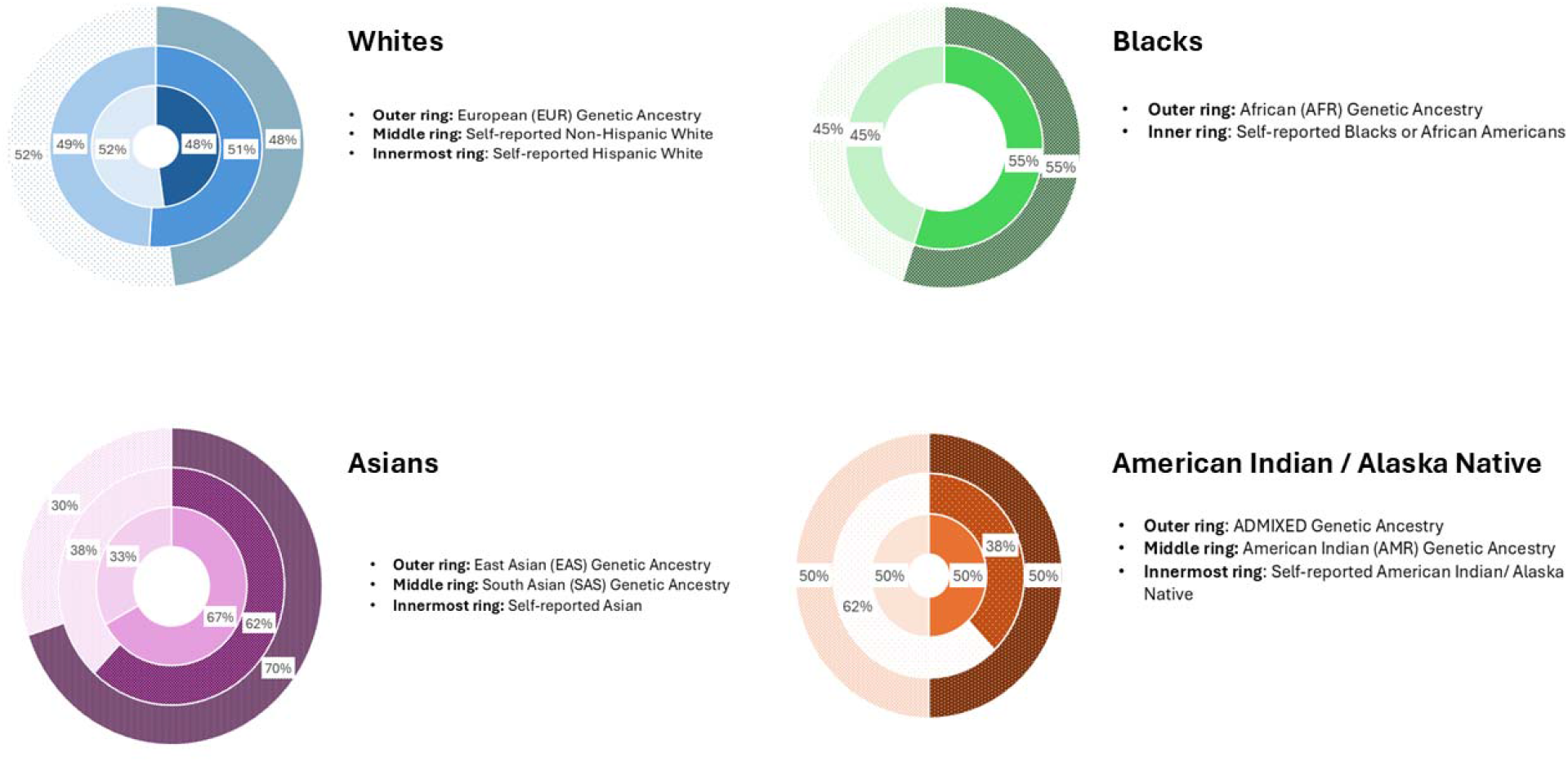
Prevalence of Early Psychosis in AD when using self-reported ancestry vs. genetic ancestry. For each ring, the darker color represents the prevalence of having the symptom, while the lighter color represents the prevalence of not having the symptom.

**Figure 6.**
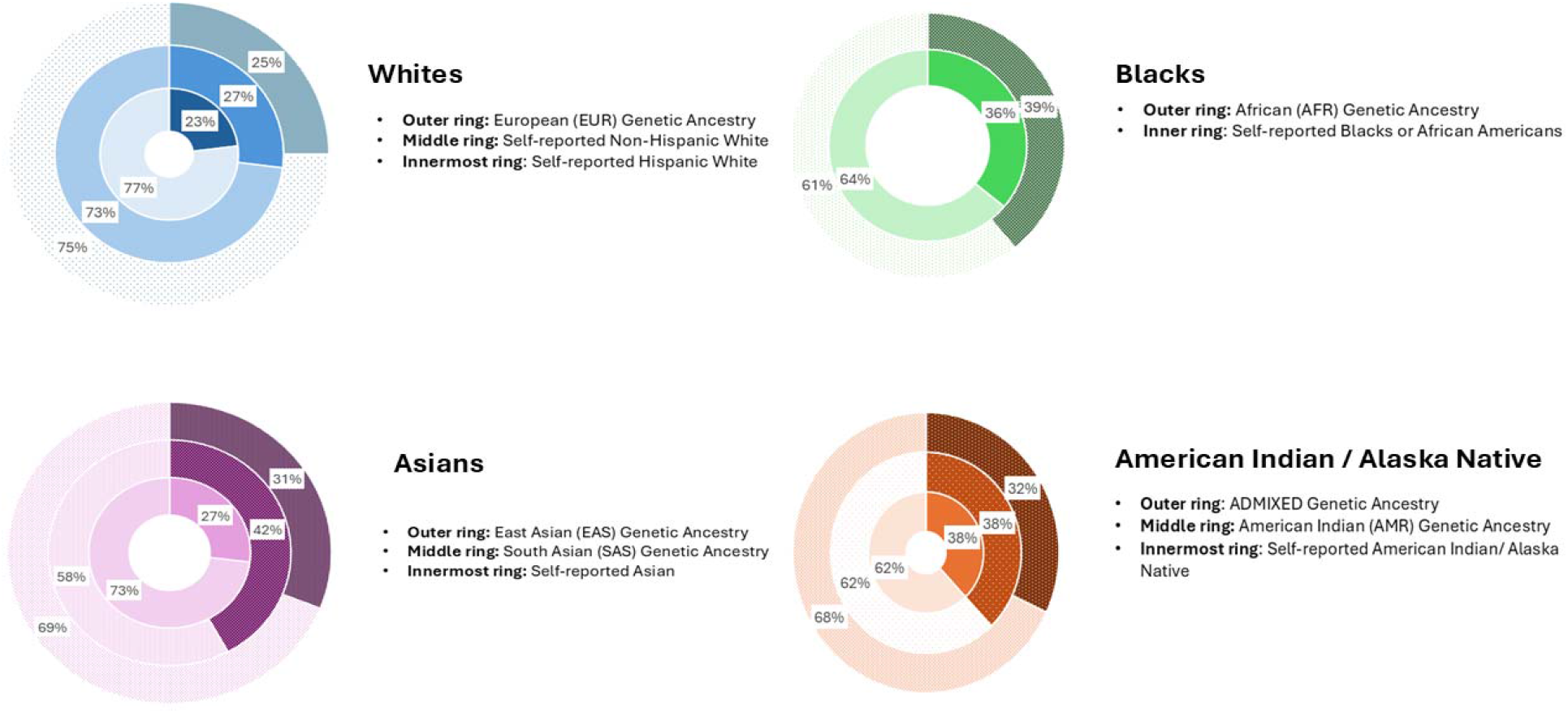
Prevalence of Late Psychosis in AD when using self-reported ancestry vs. genetic ancestry. For each ring, the darker color represents the prevalence of having the symptom, while the lighter color represents the prevalence of not having the symptom.

**Figure 7.**
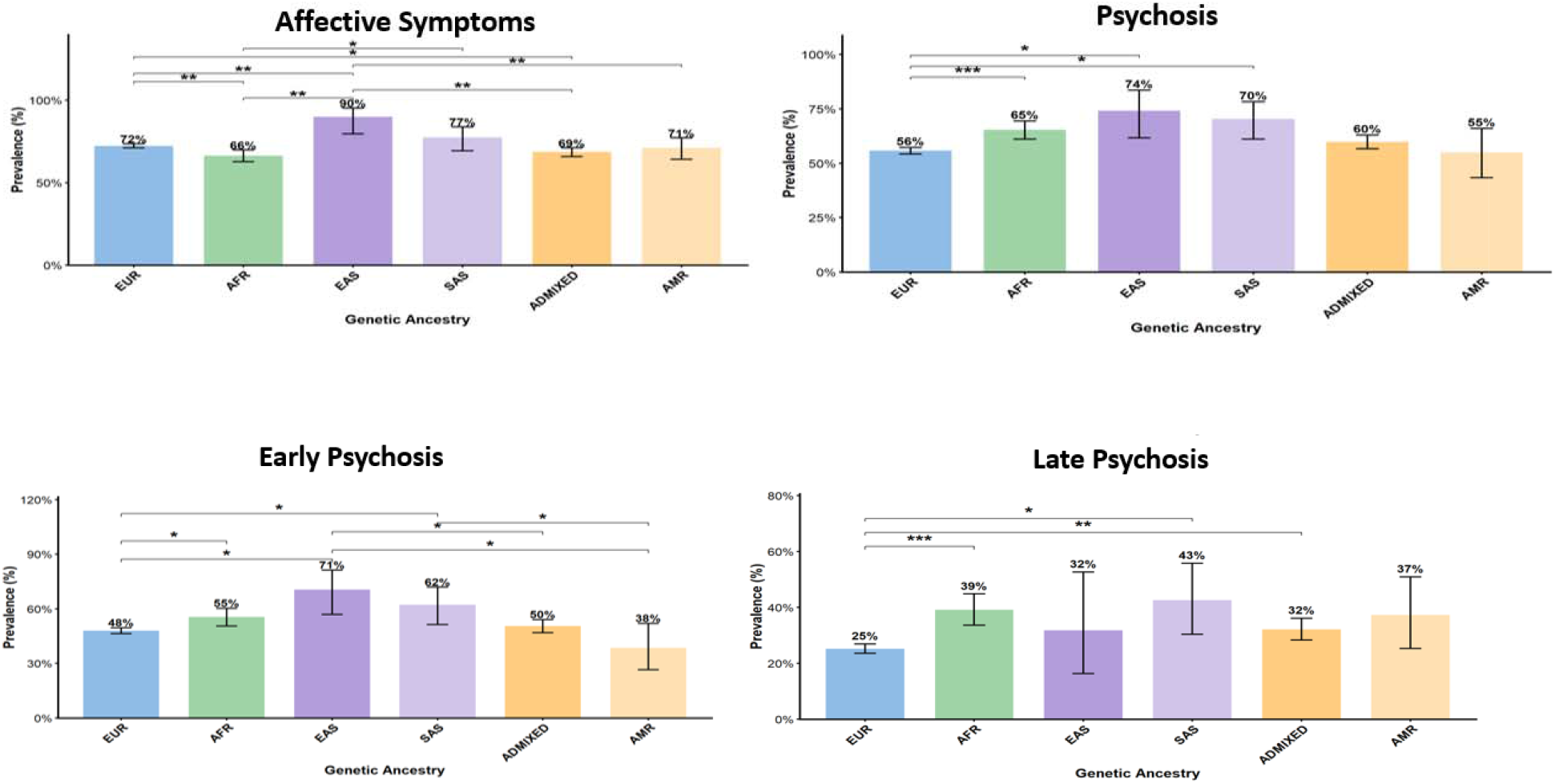
Comparison of Prevalence of Affective Symptoms, Psychosis, Early Psychosis, and Late Psychosis in AD across genetic ancestry groups. EUR=European ancestry; AFR=African ancestry; SAS=South Asian ancestry; EAS=East Asian ancestry; ADMIXED=admixed genetic ancestry; AMR=Amerindian ancestry. The horizontal bars represent the level of significance between two ancestry groups. Level of Significance: * = p < 0.05, ** = p < 0.01, *** = p <0.001.

## DISCUSSION

Studies of group differences in NPS have historically relied on self-reported race and ethnicity. While these measures are essential for capturing social context and structural inequities, they are not designed to represent biological inherited genetic variation. Our findings suggest that genetic ancestry and self-reported race/ethnicity capture overlapping but non-equivalent dimensions of population structure, with important implications for interpreting NPS risk in AD. The high concordance observed for individuals self-identifying as Black and clustering with AFR ancestry suggests that, in this group, self-reported ethnicity may reasonably approximate underlying genetic ancestry. In contrast, the substantial heterogeneity observed across most other groups— particularly among Hispanic White, Asian, and American Indian/Alaska Native individuals— underscores the limitations of self-reported categories in genetically admixed or diverse populations.

This distinction is especially relevant for affective symptoms, where differences between self-reported race/ethnicity and genetic ancestry were more pronounced and consistently statistically significant. The systematically lower prevalence of affective symptoms in genetically defined EUR, ADMIXED, AMR, and SAS groups compared to their corresponding self-reported categories suggests that reliance on self-reported race/ethnicity alone may overestimate or obscure underlying biological heterogeneity [25–27]. These discrepancies likely reflect the combined influence of genetic background and non-genetic factors embedded within self-reported categories, including sociocultural context, environmental exposures, healthcare access, diagnostic practices, and race– or ethnicity-associated diagnostic bias in clinical assessment [28–30]. The particularly high prevalence observed in EAS and SAS ancestry groups further points to potential ancestry-linked biological or gene–environment contributors to affective symptom vulnerability that warrant deeper investigation.

In contrast, psychosis showed smaller and less consistent differences between self-reported race/ethnicity and genetic ancestry, with only modest but statistically significant differences observed for EUR versus non-Hispanic White individuals. While we cannot exclude the possibility that some of the comparisons on psychosis were statistically underpowered, these observations may suggest that psychosis in AD may be less sensitive to ancestry-related stratification, or more strongly influenced by other non-ancestry-related factors. The observation that psychosis prevalence was lowest in EUR and AMR rather than AFR—unlike affective symptoms—further supports the notion that different NPS domains have at least partially distinct etiological architectures. The smaller discrepancy between self-reported and genetic ancestry for psychotic than affective symptoms is of note, as psychosis is often thought of as a more biologically based NPS than affective symptoms [31, 32].

Taken together, these findings indicate that genetic ancestry offers complementary and more refined resolution of NPS heterogeneity than self-reported race/ethnicity alone. This has important implications for precision medicine. Neuropsychiatric symptoms in AD are clinically heterogeneous and frequently exhibit variable treatment responses [33], and identifying ancestry-related differences in symptom profiles may help define biologically meaningful patient subtypes, improve gene–symptom mapping, and inform hypotheses regarding underlying molecular mechanisms. Including diverse populations and incorporating genetic ancestry into NPS research may also enhance individualized risk prediction and support the development of more targeted therapeutic approaches. Beyond clinical translation, these results carry broader public health relevance. NPS are key drivers of caregiver burden, healthcare utilization, and institutionalization [34]. Improved characterization of NPS prevalence across ancestry groups may enable more accurate identification of populations at elevated risk for specific symptom profiles, thereby informing screening strategies, caregiver support, and the allocation of behavioral health resources in increasingly diverse aging populations.

## RESEARCH IN CONTEXT

1. **Systematic Review:** Previous studies have documented racial and ethnic differences in neuropsychiatric symptoms (NPS) among individuals with Alzheimer’s disease (AD). However, most investigations have relied on self-reported race and ethnicity as proxies for biological variation. While these constructs are essential for understanding social determinants of health, they do not directly capture genetic background. Few studies have examined how genetically inferred ancestry relates to the prevalence of NPS in AD, and direct comparisons between self-reported race and genetic ancestry in this context are limited.
2. **Interpretation:** In a large, diverse cohort from the Alzheimer’s Disease Sequencing Project, we found that estimates of NPS prevalence differed depending on whether population structure was defined using self-reported race or genetically inferred ancestry. We also observed significant differences in the prevalence of individual NPS across genetically defined ancestry groups. These findings suggest that genetic ancestry captures biologically relevant heterogeneity in neuropsychiatric symptom burden that is not fully reflected by socially defined racial categories, supporting its value for refining phenotypes and advancing precision-medicine approaches in AD.
3. **Future Directions:** Future research should integrate genetic ancestry with social determinants of health, longitudinal symptom trajectories, and multi-omic data to better disentangle biological and environmental contributions to NPS in AD. Expanding representation of diverse and admixed populations in genetic studies will be critical to ensure equitable advances in risk prediction, biomarker discovery, and development of targeted interventions for neuropsychiatric symptoms.

## HIGHLIGHTS

- Neuropsychiatric symptom (NPS) prevalence in Alzheimer’s disease differs when defined by self-reported race versus genetically inferred ancestry.
- Significant variation in individual NPS was observed across genetically defined ancestry groups.
- Genetic ancestry captures biologically relevant heterogeneity not fully reflected by racial categories.
- Incorporating genetic ancestry refines phenotypic characterization and supports precision-medicine approaches to NPS.
- Ancestry-informed prevalence estimates also have public health value for targeted screening, care planning, and resource allocation.

## CONSENT STATEMENT

This study utilized only de-identified, clinical, demographic and whole-genome sequencing data. No new human subjects were recruited or directly studied. All original studies from which these data were derived obtained informed consent from participants and received approval from their respective institutional review boards (IRBs). Therefore, additional informed consent was not required for this secondary analysis.

## Data Availability

The data used in this study are available through the National Institute on Aging Genetics of Alzheimer's Disease Data Storage Site (NIAGADS) Data Sharing Service. Access to individual-level Alzheimer's Disease Sequencing Project (ADSP) data is controlled and requires an approved data access request in accordance with NIAGADS and NIH data-use policies.

https://dss.niagads.org/datasets/ng00067/

