## Supplemental Material for "Discordance Between Genetic Ancestry and Self-Reported Race Impacts Inference of Neuropsychiatric Burden in Alzheimer’s Disease"

### **Supplementary Table 1.** Sample Transitions between Self -Reported Race/Ethnicity and Genetic Ancestry Groups

| **Self-Reported Race/Ethnicity** | | **Transition to genetically defined ancestry cluster (n (%))** | | | | | |
| --- | --- | --- | --- | --- | --- | --- | --- |
|  | **Total** | **ADMIXED** | **AMR** | **EUR** | **SAS** | **AFR** | **EAS** |
| Non-Hispanic White | 4451 | 16 (0.36%) | 0 (0%) | 4429 (99%) | 2 (0.04%) | 4 (0.1%) | 0 (0%) |
| Hispanic White | 688 | 467 (68%) | 5 (0.7%) | 134 (19.5%) | 79 (11.5%) | 3 (0.4%) | 0 (0%) |
| White | 257 | 0 (0%) | 0 (0%) | 255 (99%) | 0 (0%) | 2 (0.8%) | 0 (0%) |
| Black or African American | 805 | 183 (23%) | 0 (0%) | 2 (0.25%) | 1 (0.12%) | 619 (77%) | 0 (0%) |
| Asian | 75 | 8 (10.7%) | 0 (0%) | 1 (1%) | 10 (13.4%) | 1 (1.3%) | 55 (73%) |
| American Indian/Alaska Native | 46 | 19 (41.3%) | 8 (17.4%) | 17 (37%) | 2 (4.4%) | 0 (0%) | 0 (0%) |
| NA | 270 | 133 (49%) | 112 (41.5%) | 1 (0.4%) | 20 (7%) | 1 (0.4%) | 3 (1%) |
| Other | 526 | 367 (70%) | 58 (11%) | 31 (6%) | 10 (2%) | 59 (11%) | 1 (0.2%) |
| Total | 7118 | 1193 (17%) | 183 (2.6%) | 4870 (68%) | 124 (1.8%) | 689 (9.6%) | 59 (0.8%) |

ADMIXED, genetically admixed ancestry; AMR, American Indian genetic ancestry; EUR, European genetic ancestry; SAS, South Asian genetic ancestry; AFR, African genetic ancestry; EAS, East Asian genetic ancestry; NA, self-reported race/ethnicity not available.

### **Supplemental Table 2.** Comparison of NPS prevalence between self-reported race and genetic ancestry using Fisher’s exact test

| **Genetic ancestry / Self-reported Race** | **Affective Symptoms** | | **Psychosis** | | **Early Psychosis** | | **Late Psychosis** | |
| --- | --- | --- | --- | --- | --- | --- | --- | --- |
|  | **prevalence** | **p-value** | **prevalence** | **p-value** | **prevalence** | **p-value** | **prevalence** | **p-value** |
| **EUR vs.**  **Self-Reported White** |  |  |  |  |  |  |  |  |
| EUR | 72% | REF | 56% | REF | 48% | REF | 25% | REF |
| Non-Hispanic White | 76% | **1.45 × 10⁻⁵** | 59% | **0.002** | 51% | **0.001** | **27%** | **0.03** |
| Hispanic White | 62% | **8.39 × 10⁻⁸** | 53% | 0.21 | 48% | 0.93 | 23% | 0.46 |
| **AFR vs.**  **Self-Reported Black** |  |  |  |  |  |  |  |  |
| AFR | 66% | 0.78 | 65% | 0.75 | 55% | 0.94 | 39% | 0.57 |
| African American | 67% | REF | 64% | REF | 55% | REF | 36% | REF |
| **EAS, SAS vs.**  **Self-Reported Asian** |  |  |  |  |  |  |  |  |
| EAS | 90% | 0.79 | 74% | 0.56 | 70% | 0.70 | 31% | 0.76 |
| SAS | 77% | 0.08 | 70% | 0.74 | 62% | 0.62 | 42% | 0.17 |
| Asian | 88% | REF | 68% | REF | 67% | REF | 27% | REF |
| **ADMIXED, AMR vs.**  **Self-Reported American Indian/Alaska Native** |  |  |  |  |  |  |  |  |
| ADMIXED | 69% | **0.008** | 60% | 0.53 | 50% | 1.00 | 32% | 0.52 |
| AMR | 71% | **0.03** | 55% | 0.33 | 38% | 0.24 | 38% | 1.00 |
| American Indian/Alaska Native | 87% | REF | 65% | REF | 50% | REF | 38% | REF |

ADMIXED, genetically admixed ancestry; AMR, American Indian genetic ancestry; EUR, European genetic ancestry; SAS, South Asian genetic ancestry; AFR, African genetic ancestry; EAS, East Asian genetic ancestry.

### **Supplementary Figure 1.** Agreement between self-reported race and genetically inferred ancestry clusters estimated using Cohen’s kappa coefficient (κ). Points represent κ values for each race–ancestry comparison, and colors indicate the strength of agreement according to the Landis and Koch classification scheme. Higher positive κ values indicate stronger agreement, lower positive κ values indicate slight agreement, whereas negative values indicate disagreement. ADMIXED, genetically admixed ancestry; AMR, Admixed American ancestry; EUR, European ancestry; SAS, South Asian ancestry; AFR, African ancestry; EAS, East Asian ancestry.

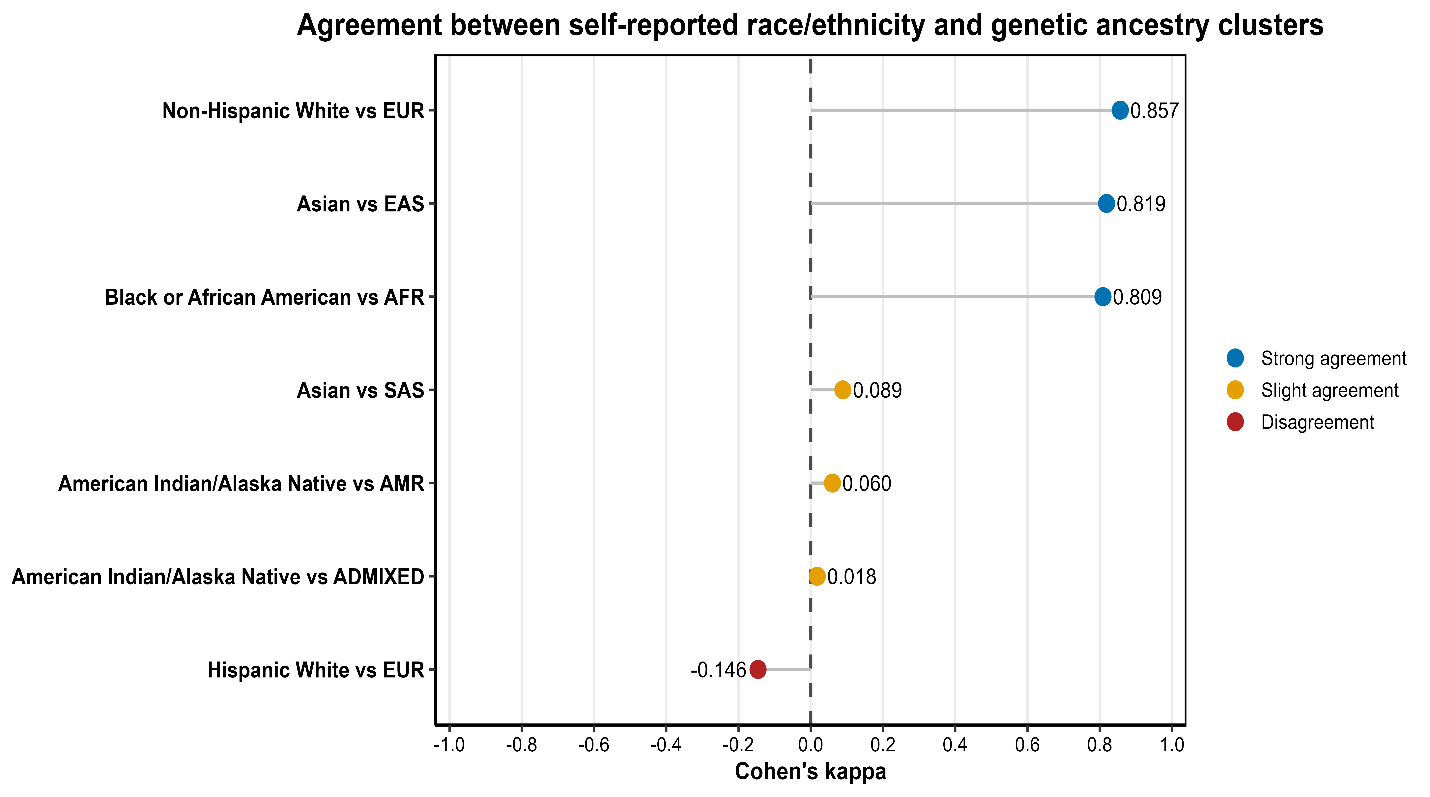
